# Diagnosis of left atrial appendage thrombus using cardiac computed tomography: new insights from thrombi locations

**DOI:** 10.1101/2023.02.24.23286435

**Authors:** Chuxian Guo, Zhi Jiang, Jionghong He, Haiyan Ma, Yuquan Wang, Jing Tan, Qiaoqiao Ou, Ye Tian, Longhai Tian, Qifang Liu, Jing Huang, Long Yang

## Abstract

**Background:** Cardiac computed tomography (CCT) is an emerging non-invasive modality for assessing left atrial appendage (LAA) thrombus, but the results were conflicting. Our study aims to evaluate the accuracy of CCT for detecting LAA thrombus in patients undergoing catheter ablation of atrial fibrillation (AF), using transesophageal echocardiography (TEE) as the reference standard.

**Methods:** From May 2017 to December 2022, 726 patients (male: 60.2%, age: 61±11 years) who had both TEE and CCT before catheter ablation of AF were retrospectively included. The CCT protocol consisted of one angiographic phase and one delayed scan 30 seconds later. LAA thrombi were defined as solid masses on TEE or persistent defects on CCT. The thrombus dimension and location, the LAA filling and emptying flow velocity were assessed by TEE.

**Results:** Of the 57(7.9%) patients with LAA thrombi identified by TEE, 29(50.9%) were located at the LAA ostium, and 28(49.1%) were in the LAA. The former showed higher motility following blood flow and heartbeats than the latter. The CCT detected 14(48.3%) of the LAA-ostium thrombi but 25(89.3%) of those in the LAA (*p* = 0.001). The LAA-ostium thrombi with the LAA mean flow velocity higher than 0.35m/s and maximum diameters shorter than 10mm were more prone to have CCT false-negative results.

**Conclusion:** For patients undergoing catheter ablation for AF, CCT with a 30s delay scan is less sensitive to LAA thrombi than TEE, especially LAA-ostium thrombi with smaller sizes and higher LAA flow velocity.

**Clinical perspective section:** 

**What are new?:**

1. Over half of the LAA thrombi were located at the LAA ostium.
2. The CCT was less sensitive to the LAA-ostium thrombi with smaller sizes and higher LAA flow velocity.

**What is the clinical implication?:** 1. The CCT using a 30s delay scan did not reliably exclude the LAA thrombi for the patients scheduled for pulmonary vein isolation, especially those located at the LAA ostium.

## Introduction

Transesophageal echocardiology (TEE) is currently the gold standard for the assessment of left atrial appendage (LAA) thrombi in patients undergoing catheter ablation of atrial fibrillation (AF)^1^. In the past decades, cardiac computed tomography (CCT) has been widely studied for detecting LAA thrombus^2-5^. Recently, the delayed contrast-enhanced CCT at 6 minutes was reported to reach 100% sensitivity and negative predictive value in detecting LAA thrombus^6^. For patients who are not tolerant of the discomfort associated with TEE, CCT is proposed as a reliable alternative^7^. However, the accuracy of CCT varied between studies^2-5, 8^, raising the safety concern and limiting the routine use of CCT.

The left atrial thrombi were classified into a movable ball, fixed ball, and mountain according to their morphology^9^. Around 1/3 of the thrombi in the left atrium were movable. Although CCT is of high spatial resolution, its temporal resolution is 4∼17Hz, and it takes 60ms ∼ 250ms to acquire one slice^8, 10-12^. Compared to TEE, which allows live display with a high spatiotemporal resolution (<1mm, 20∼30Hz)^13^, CCT is theoretically inferior for identifying moving objects in the left atrium. In addition, the circulatory stasis and the fibrillation of the atrial wall during AF also increase the difficulty of detecting movable thrombi^14^. In previous studies, the presence of LAA thrombus was 2(0.24%) ∼16(19.04%)^2-5,15^. The high sensitivity and negative predictive value of CCT could be attributed to the low incidence of movable thrombus. Therefore, it is essential to investigate the diagnostic accuracy of CCT in a larger cohort with a higher incidence of LAA thrombus.

In the current study, we retrospectively included 726 patients who had both TEE and CCT before catheter ablation for AF. 57(7.9%) patients were diagnosed with LAA thrombi using TEE. We aimed to evaluate the sensitivity of CCT in detecting LAA thrombus and to analyze the risk factors of false-negative results.

## Methods

### Study population

From May 2017 to December 2022, patients were routinely scheduled for TEE and CCT before catheter ablation of AF in Guizhou Provincial People’s Hospital. Those who had both examinations within a 12-hour interval on the same day were retrospectively included. All the patient characteristics and images were acquired from the clinical database. The CHA_2_DS_2_-VASc score was calculated following the 2020 ESC guideline^1^. The study was approved by the institutional ethics committee of Guizhou Provincial People’s Hospital. The informed consent was waived due to the retrospective nature, and no identified information was collected.

### CCT protocol

All patients were examined with a third-generation dual-source CT (SOMATOM Force, Siemens Healthineers, Forchheim, Germany). The imaging protocol included a standard angiographic-phase acquisition to assess the anatomy of the left atrium and one delayed-phase acquisition at 30 s after contrast injection to detect thrombi. No electrocardiography gating was used. The tube voltage and current were automatically adapted using the CARE kV or CARE DOSE 4D with the reference of 100kv and 250mA. The gantry rotation time was 250ms, and the detector collimator was 192×0.6 mm. The scanning image covers the range from the aortic arch to the cardiac base. The monitoring site was localized within the left atrium. The contrast material containing 350 mg/ml of iodine was administrated intravenously at 4-5 ml/s with a total dose of 35 ml by a double-barrel syringe, followed by the same amount of 0.9% saline. For the angiographic phase, the scanning was triggered when the monitoring site reached 90 HU. Then the delayed phase was acquired 30 s later. All images were reconstructed using the ADMIRE algorithm grade 3 with a layer thickness of 1 mm and interlayer spacing of 0.7 mm. The 3D left atrium and pulmonary veins were reconstructed using the Syngo.via software (Siemens Healthcare, Forchheim, Germany).

### CCT image analysis

Images were transferred to an external workstation (Syngo VPCT body, Siemens Healthcare, Forchheim, Germany). Two independent radiologists with more than eight years of cardiovascular imaging, blinded to all patients’ data, retrospectively evaluated the presence of LAA thrombus and LAA morphology. Thrombus was defined as a persistent filling defect in the 30s-delayed scans. The LAA morphology was classified into simple or complex according to their main and branching structures on the 3D reconstructions^16^. In disagreement, the consensus was achieved in a final joint reading.

### Transesophageal echocardiology

A PhilipsiEPIQ7C color Doppler ultrasound diagnostic instrument (Philips, Amsterdam, the Netherlands) and an X7-2t probe were used. The left atrium and LAA were scanned using a complete 2D, colored, pulsed, and continuous-wave Doppler echocardiogram according to the EACVI recommendations^17^. Thrombi were defined as circumscribed echogenic or echolucent masses distinct from the atrial wall. LAA ostium was defined as within the 5mm range of the junction between the smooth atrial wall and trabeculated LAA. The dimensions and locations of thrombi were assessed if present. The LAA filling and emptying velocity was averaged by ten consecutive measurements of the backward and forward flow rates using the pulsed wave Doppler. LAA mean flow velocity (LAAMFV) was calculated by halving the sum of the LAA filling and emptying velocity. Two independent cardiologists with more than five years of experience in echocardiology evaluated the presence of LAA thrombus and measured the flow velocity. Disagreements were resolved by consensus.

### Statistical analysis

Continuous variables were expressed as mean ± SD deviation. The Student’s t-test was used for comparison if normally distributed; otherwise, the Mann-Whitney U test was used. Categorical variables were expressed as frequency (percentage). The Chi-square test was used for comparison if the expected frequency is more than 5; otherwise, Fisher’s exact test was used. A two-tailed *p* value of less than 0.05 was considered statistically significant.

Using TEE as the reference standard, the diagnostic performance of CCT for LAA thrombi was calculated. All statistical analysis was performed using SPSS 26.0.

## Results

### Patient characteristics

A total of 726 patients (age: 61±11 years; male gender: 60.2%) with 57(7.9%) LAA thrombi diagnosed by TEE were included in the study (Figure 1). The patients with LAA thrombus had a higher incidence of previous stroke/transient ischemic attack (TIA), larger left atrial size, worse left ventricular function and a higher rate of complex LAA morphology than those without LAA thrombus (*p* < 0.05) (Table 1). 29(50.9%) of the thrombi were located at the LAA ostium and 28(49.1%) were in the LAA. The clinical characteristics and maximum thrombus diameter were similar between the patients. The thrombi at the LAA ostium were accompanied by higher LAA filling and emptying velocities than those in the LAA (Table 2). The former also showed significantly higher mobility following the blood flow and cardiac motion than the latter (Supplementary material online, Video S1).

**Table 1.**
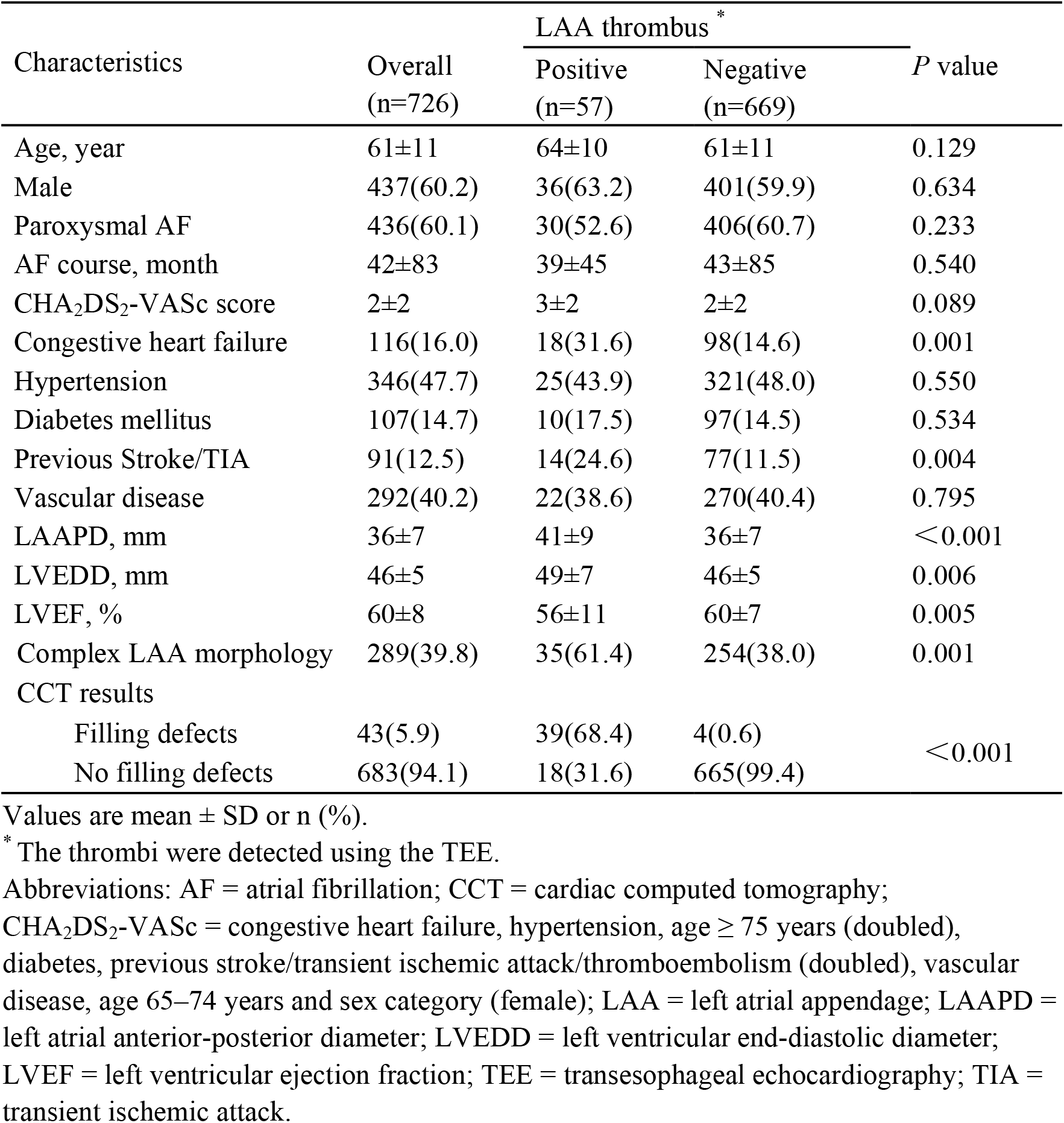
Patient characteristics.

| Characteristics | Overall<br>(n=726) | LAA thrombus * |  | *P* value |
| --- | --- | --- | --- | --- |
|  |  | Positive<br>(n=57) | Negative<br>(n=669) |  |
| Age, year | 61±11 | 64±10 | 61±11 | 0.129 |
| Male | 437(60.2) | 36(63.2) | 401(59.9) | 0.634 |
| Paroxysmal AF | 436(60.1) | 30(52.6) | 406(60.7) | 0.233 |
| AF course, month | 42±83 | 39±45 | 43±85 | 0.540 |
| CHA<sub>2</sub>DS<sub>2</sub>-VASc score | 2±2 | 3±2 | 2±2 | 0.089 |
| Congestive heart failure | 116(16.0) | 18(31.6) | 98(14.6) | 0.001 |
| Hypertension | 346(47.7) | 25(43.9) | 321(48.0) | 0.550 |
| Diabetes mellitus | 107(14.7) | 10(17.5) | 97(14.5) | 0.534 |
| Previous Stroke/TIA | 91(12.5) | 14(24.6) | 77(11.5) | 0.004 |
| Vascular disease | 292(40.2) | 22(38.6) | 270(40.4) | 0.795 |
| LAAPD, mm | 36±7 | 41±9 | 36±7 | <0.001 |
| LVEDD, mm | 46±5 | 49±7 | 46±5 | 0.006 |
| LVEF, % | 60±8 | 56±11 | 60±7 | 0.005 |
| Complex LAA morphology | 289(39.8) | 35(61.4) | 254(38.0) | 0.001 |
| CCT results |  |  |  |  |
| Filling defects | 43(5.9) | 39(68.4) | 4(0.6) | <0.001 |
| No filling defects | 683(94.1) | 18(31.6) | 665(99.4) |  |

Values are mean ± SD or n (%).
\* The thrombi were detected using the TEE.
Abbreviations: AF = atrial fibrillation; CCT = cardiac computed tomography; CHA<sub>2</sub>DS<sub>2</sub>-VASc = congestive heart failure, hypertension, age ≥ 75 years (doubled), diabetes, previous stroke/transient ischemic attack/thromboembolism (doubled), vascular disease, age 65–74 years and sex category (female); LAA = left atrial appendage; LAAPD = left atrial anterior-posterior diameter; LVEDD = left ventricular end-diastolic diameter; LVEF = left ventricular ejection fraction; TEE = transesophageal echocardiography; TIA = transient ischemic attack.

**Table 2.** Patient characteristics between thrombus locations.

| Characteristics | Thrombus location * |  | *P* value |
| --- | --- | --- | --- |
|  | LAA ostium (n=29) | In LAA(n=28) |  |
| Age, years | 63±10 | 64±10 | 0.886 |
| Male | 20(69.0) | 16(57.1) | 0.355 |
| Paroxysmal AF | 18(62.1) | 12(42.9) | 0.146 |
| AF course, month | 40±52 | 39±35 | 0.648 |
| CHA<sub>2</sub>DS<sub>2</sub>-VASc score | 2±2 | 3±2 | 0.220 |
| Congestive heart failure | 9(31.0) | 9(32.1) | 0.928 |
| Hypertension | 10(34.5) | 15(53.6) | 0.147 |
| Diabetes mellitus | 5(17.2) | 5(17.9) | 1.000 |
| Previous Stroke/TIA | 7(24.1) | 7(25.0) | 0.940 |
| Vascular disease | 9(31.0) | 13(46.4) | 0.233 |
| LAAPD, mm | 41±10 | 40±8 | 0.873 |
| LVEDD, mm | 49±6 | 49±8 | 0.492 |
| LVEF, % | 57±11 | 55±11 | 0.424 |
| Complex LAA | 17(58.6) | 18(64.3) | 0.661 |
| LAA filling velocity, m/s | 0.48±0.19 | 0.37±0.19 | 0.021 |
| LAA emptying velocity, m/s | 0.47±0.20 | 0.38±0.16 | 0.050 |
| Thrombus maximum diameter, mm | 10±4 | 14±10 | 0.045 |
| CCT results |  |  |  |
| Filling defects | 14(48.3) | 25(89.3) | 0.001 |
| No filling defects | 15(51.7) | 3(10.7) |  |

Values are mean ± SD or n (%).
\* The thrombus locations were determined using the TEE.
Abbreviations: AF = atrial fibrillation; CCT = cardiac computed tomography;
CHA<sub>2</sub>DS<sub>2</sub>-VASc = congestive heart failure, hypertension, age ≥ 75 years (doubled),
diabetes, previous stroke/transient ischemic attack/thromboembolism (doubled), vascular
disease, age 65–74 years and sex category (female); LAA = left atrial appendage; LAAPD=
left atrial anterior-posterior diameter; LVEDD = left ventricular end-diastolic diameter;
LVEF = left ventricular ejection fraction; TEE = transesophageal echocardiography; TIA =
transient ischemic attack.

**Figure 1.**
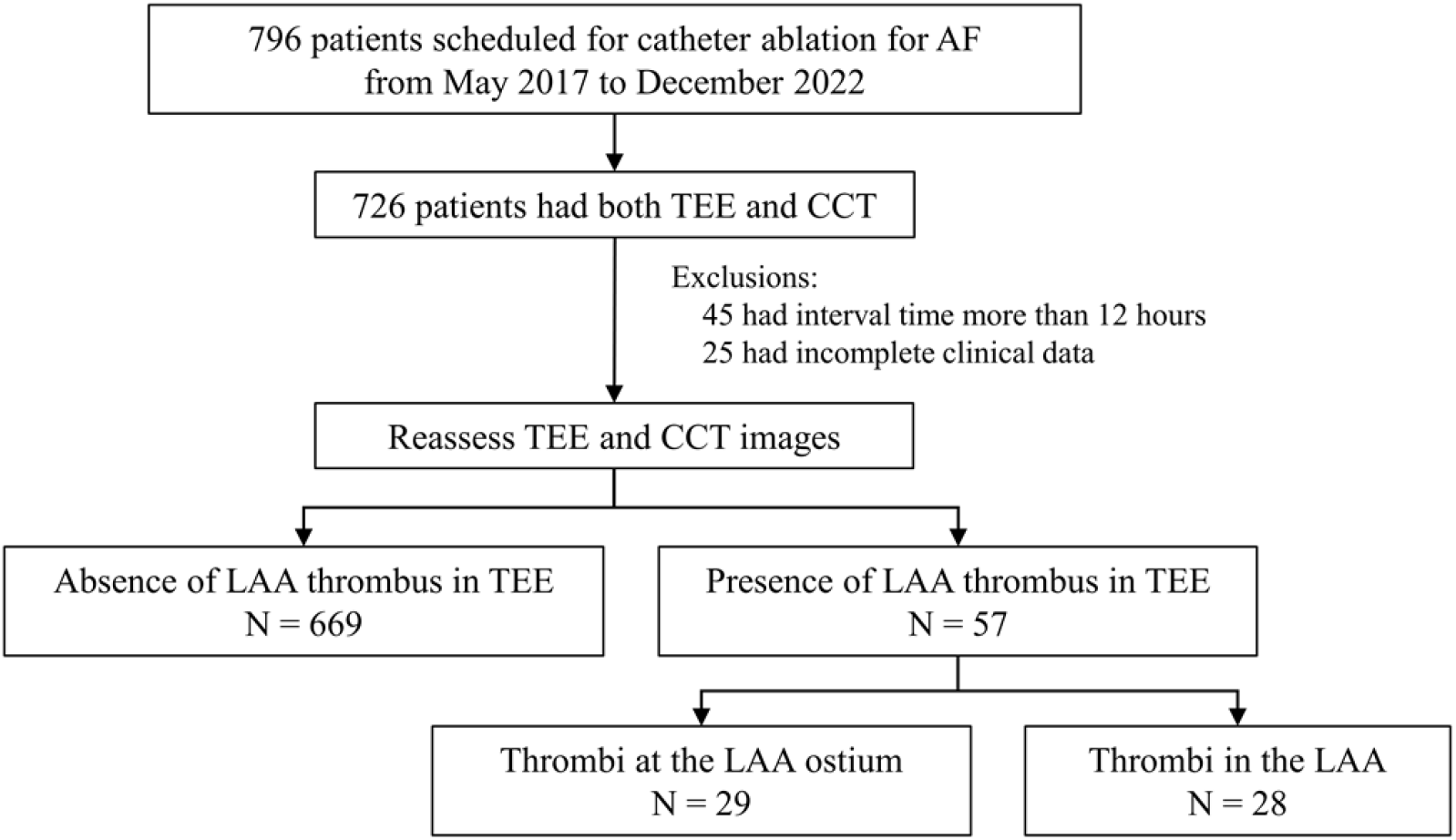
Patient flowchart. **Abbreviations:** AF = atrial fibrillation; CCT = cardiac computed tomography; LAA = left atrial appendage; TEE = transesophageal echocardiography.

### Diagnostic accuracy of CCT

The CCT had an overall sensitivity of 68.4% and a specificity of 99.4% to the LAA thrombi (Table 1). However, the CCT was less sensitive to the thrombi at LAA ostium than those in the LAA (48.3% vs. 89.3%, *p* = 0.001) (Table 2). Representative images are shown in Figure 2.

**Figure 2.**
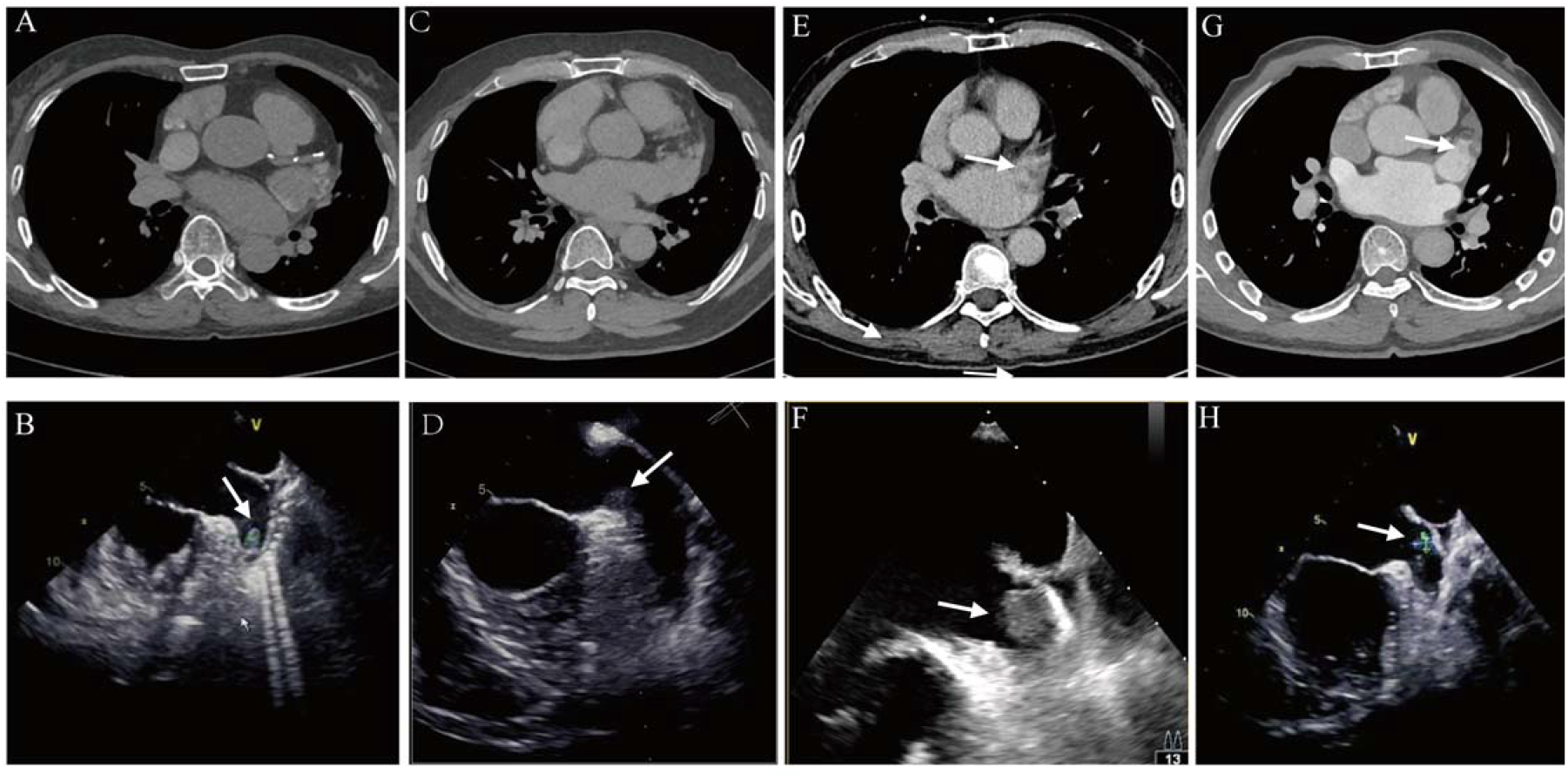
Representative TEE and CCT images of the LAA thrombi. The upper panel shows CCT images. The lower panel shows TEE images. A-B. Representative false-negative CCT result. The thrombus was located in the LAA. C-D. Representative false-negative CCT result. The thrombus was located at the LAA ostium. E-F. Representative true-positive CCT result. The thrombus was located in the LAA. G-H. Representative true-positive CCT result. The thrombus was located at the LAA ostium. White arrows denote the thrombi.

Given that high sensitivity is essential to exclude LAA thrombus, the CCT results of the LAA thrombi underwent further analysis. Compared with the CCT true-positive thrombi, the false-negative ones more frequently located at the LAA ostium (83.3% vs. 35.9%, *p* = 0.001), were shorter in the maximum diameters (8±2mm vs. 14±9mm, *p* < 0.001), and were accompanied by higher LAA filling and emptying velocities (0.53±0.15 m/s vs. 0.38±0.19 m/s, *p* = 0.001; 0.55±0.21 m/s vs. 0.37±0.14 m/s, *p* < 0.001) (Table 3). By scattering the CCT results on the 2-D plane that consisted of LAAMFV and thrombus maximum diameter, the thrombi at the LAA ostium with the LAAMFVs higher than 0.35m/s and maximum diameters shorter than 10mm were more prone to have CCT false-negative results (Figure 3A). The CCT false-negative thrombi in the LAA had maximum diameters shorter than 8mm (Figure 3B).

**Table 3.** Patient characteristics between CCT true-positive and false-negative results.

| Characteristics | LAA thrombus * |  | *P* value |
| --- | --- | --- | --- |
|  | CCT false-negative result<br>(n=18) | CCT true-positive result<br>(n=39) |  |
| Age, years | 64±9 | 64±10 | 0.810 |
| Male | 11(61.1) | 25(64.1) | 0.828 |
| Paroxysmal AF | 12(66.7) | 18(46.2) | 0.149 |
| AF course, month | 42±54 | 38±40 | 0.952 |
| CHA<sub>2</sub>DS<sub>2</sub>-VASc score | 3±2 | 3±2 | 0.450 |
| Congestive heart failure | 5(27.8) | 13(33.3) | 0.675 |
| Hypertension | 10(55.6) | 15(38.5) | 0.227 |
| Diabetes mellitus | 5(27.8) | 5(12.8) | 0.168 |
| Previous Stroke/TIA | 5(27.8) | 9(23.1) | 0.702 |
| Vascular disease | 7(38.9) | 15(38.5) | 0.975 |
| LAAPD, mm | 41±9 | 41±9 | 0.770 |
| LVEDD, mm | 49±6 | 49±7 | 0.336 |
| LVEF, % | 59±9 | 55±12 | 0.331 |
| Complex LAA | 9(50.0) | 26(66.7) | 0.230 |
| LAA filling velocity, m/s | 0.53±0.15 | 0.38±0.19 | 0.002 |
| LAA emptying velocity, m/s | 0.55±0.21 | 0.37±0.14 | 0.001 |
| Thrombi maximum diameter, mm | 8±2 | 14±9 | <0.001 |
| Thrombus location |  |  |  |
| LAA ostium | 15(83.3) | 14(35.9) | 0.001 |
| In LAA | 3(16.7) | 25(64.1) |  |

Values are mean ± SD or n (%).
\* The thrombi were detected using the TEE.
Abbreviations: AF = atrial fibrillation; CCT = cardiac computed tomography; CHA<sub>2</sub>DS<sub>2</sub>-VASc = congestive heart failure, hypertension, age ≥ 75 years (doubled), diabetes, previous stroke/transient ischemic attack/thromboembolism (doubled), vascular disease, age 65–74 years and sex category (female); LAA = left atrial appendage; LAAPD = left atrial anterior-posterior diameter; LVEDD = left ventricular end-diastolic diameter; LVEF = left ventricular ejection fraction; TEE = transesophageal echocardiography; TIA = transient ischemic attack.

**Figure 3.**
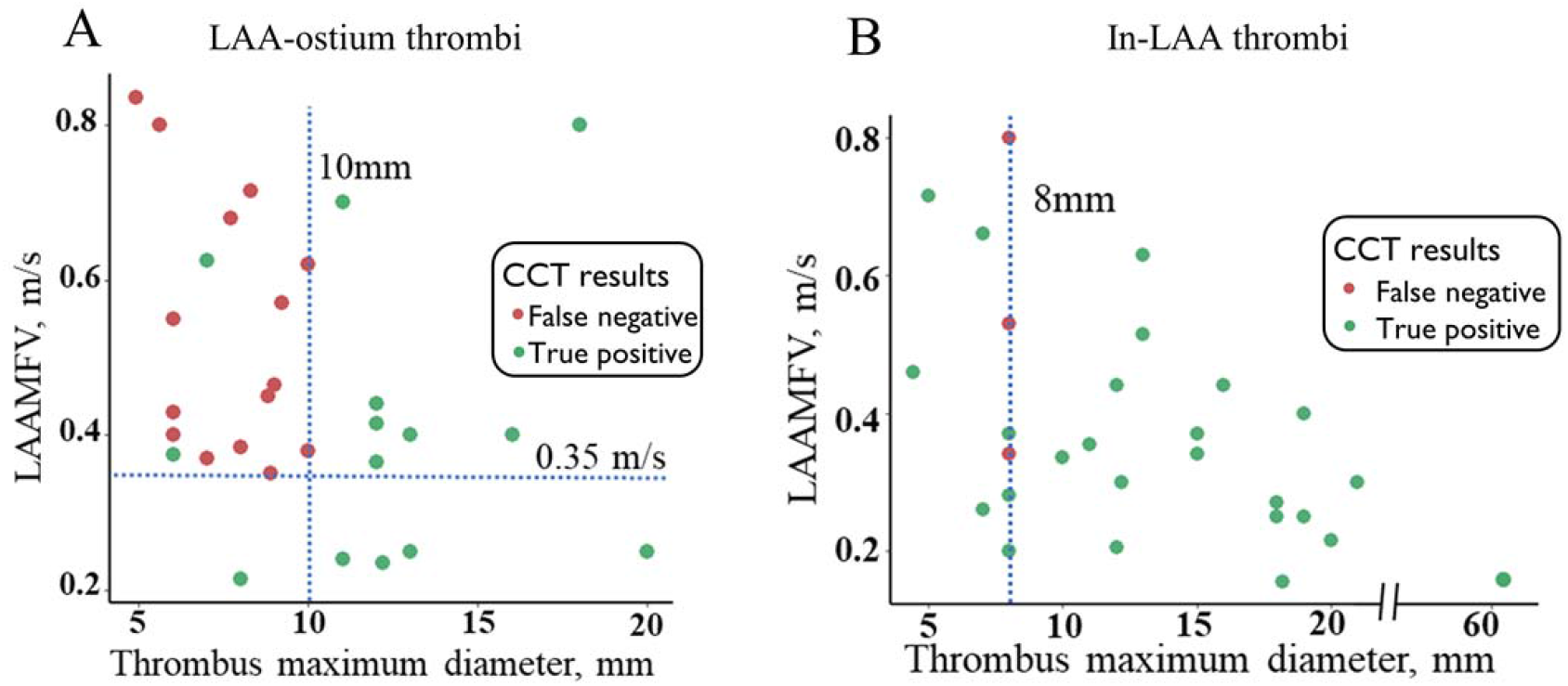
Scatter plots of CCT results of LAA thrombi. Red dots denote the CCT false-negative thrombi. Green dots denote the CCT true-positive thrombi. The LAAMFV was calculated by halving the sum of the LAA emptying and filling velocity. A. The CCT false-positive thrombi at the LAA ostium had the LAAMFV over 0.35 m/s and the maximum diameters over 10 mm. B. The CCT false-positive thrombi in the LAA ostium had a maximum diameter over 8 mm. **Abbreviations:** CCT = cardiac computed tomography; LAA = left atrial appendage; LAAMFV = left atrial appendage mean flow velocity.

## Discussion

### Major findings

We reevaluated the accuracy of CCT for detecting LAA thrombus in the 726 patients undergoing catheter ablation for AF. TEE identified 57(7.9%) patients with LAA thrombi, of which 29(50.9%) were at the LAA ostium and 28(49.1%) in the LAA. The CCT was less sensitive in detecting LAA-ostium thrombi with smaller maximum diameters and higher LAAMFV.

### Concordance

LAA thrombus is thought to form initially inside the LAA, where circulatory stasis is the greatest, and then extends towards the ostium. However, we found over half of the thrombi located at the LAA ostium. The result was concordant with a previous study that 46.3% of the thrombi were at the entrance section of LAA^9^. The presence of pits and troughs adjacent to the LAA orifice was reported in 57.7% of the human heart specimens^18^. It could be the anatomic basis for thrombosis. Another study using TEE discovered small recesses proximal to the entry of LAA in a significant number of patients, but no thrombus related to these structures was detected following LAA occlusion^19^. The discrepancy could be due to patient selection bias. Nonetheless, our findings raised concerns about the role of LAA-ostium recesses in thrombosis.

Although the third-generation 64-slice dual-source CT was employed in the current study, its sensitivity to LAA thrombi was not improved. The sensitivity of 89.3% to the thrombi in the LAA was comparable to the previous studies^2, 10, 11, 20-22^. However, the sensitivity significantly decreased for the LAA-ostium thrombi, which showed higher motility following blood flow and heartbeats than those in the LAA (Supplementary material online, Video S1). We further identified that the LAA-ostium thrombi with smaller sizes and higher LAA flow velocity were more prone to be missed using the CCT. As a higher temporal resolution is essential to detect movable objects^23^, our findings suggested the impact of thrombus motility on CCT imaging. By calculating the gantry rotation time of 250ms, the CCT had a temporal resolution of 4Hz, significantly less than that of the TEE (20∼40Hz). Thus, the low sensitivity of CCT in our study could be presumed to: (1) inadequate temporal resolution, (2) the high proportion of movable LAA-ostium thrombi, (3) the lack of ECG gating, (4) only one 30s delayed scan was used. The 4 CCT false-positive results in our study could be the misdiagnosis of thrombus due to circulatory stasis during AF^6, 24^.

### Discordance

Several studies reported that the CCT had 100% sensitivity in detecting LAA thrombi using ECG gating and single-segment reconstruction algorithm^25, 26^, or multiple delayed-phase scans^3, 14, 27^. However, these studies have the following limitations: (1) Although the single-segment reconstruction algorithm increased the CCT’s temporal resolution to 12Hz, the low incidence of LAA thrombus [2(0.24%) ∼16(19.04%)] and the less variation of thrombus location could have overestimated the sensitivity of CCT for detecting LAA thrombi in a broader range of patients. (2) The multiple delayed-phase scans distinguished the thrombi from filling defects due to circulatory stasis. Still, the technique could not necessarily increase the sensitivity to smaller and movable thrombi located at the LAA ostium. We suggest that the CCT approach to detect LAA thrombi could differ according to their location. For static thrombi in LAA, persistent filling defects in multiple delayed-phase scans have been well established. For thrombi at LAA ostium, a temporal resolution comparable to TEE and even dynamic visualization could be necessary due to their higher motility.

### Clinical implication

Our study found that the CCT with a 30s delayed scan did not reliably exclude LAA thrombus, especially those located at the LAA ostium. We did not find any clinical characteristics associated with the thrombus location. TEE is the only recommendation for stroke risk management for patients undergoing catheter ablation of AF in the 2020 ESC guideline^1^. For those who do not tolerate TEE, intracardiac echocardiography could be considered^28^.

### Limitations

Firstly, this retrospective single-center study included only patients undergoing catheter ablation for AF. The study’s findings may not be generalizable to other patients with AF. Secondly, the CCT protocol contained only one delayed-phase scan. Whether multiple delayed-phase scans reduce false-negative CCT results is unclear. Thirdly, other factors unaccounted for in this analysis – heart rhythm, atrial/ventricular rate, and blood pressure during the TEE and CCT – could affect the conclusions. Fourthly, although the interval between the TEE and CCT was less than 12 hours, it is theoretically possible for thrombi to form or disappear within this timeframe. Lastly, the incidence of LAA thrombus was higher than that in previous studies (partially due to less adherence to anticoagulation treatment), which could have affected the thrombus location and motility.

## Conclusions

In patients undergoing catheter ablation of AF, CCT with a 30s delay scan is less sensitive than TEE in detecting LAA thrombi, especially those located at LAA ostium with smaller sizes and higher LAA flow velocity.

## Data Availability

The data underlying this article may be shared upon reasonable request to the corresponding author.

## Non-standard Abbreviations and Acronyms

AF: Atrial fibrillation
CCT: Cardiac computed tomography
CHA_2_DS_2_-VASc: Congestive heart failure, hypertension, age ≥ 75 years (doubled), diabetes, previous stroke/transient ischemic attack/thromboembolism (doubled), vascular disease, age 65–74 years, and sex category (female).
LAA: Left atrial appendage
LAAMFV: Left atrial appendage mean flow velocity LAAPD Left atrial anterior-posterior diameter
LVEDD: Left ventricular end-diastolic diameter
LVEF: Left ventricular ejection fraction
TEE: Transesophageal echocardiography
TIA: Transient ischemic attack

## Acknowledgements

We thank Nicholas Tan and Jason Tri from Mayo Clinic for their dedicated help in preparing the manuscript.

## Supplementary Material

Video S1

## Supplementary Legend

**Supplementary Video1 Thrombi imaging of different location in TEE**

A-C Thrombus located at the LAA ostium. D-F Thrombus located in the LAA.

